# Intrinsic capacity as a framework for Integrated Care for Older People (ICOPE); insights from the 10/66 Dementia Research Group cohort studies in Latin America, India and China

**DOI:** 10.1101/19006403

**Authors:** Martin J. Prince, Daisy Acosta, Mariella Guerra, Yueqin Huang, KS Jacob, Ivonne Z Jimenez-Velazquez, AT Jotheeswaran, Juan J. Llibre Rodriguez, Aquiles Salas, Ana Luisa Sosa, Isaac Acosta, Rosie Mayston, Zhaorui Liu, Jorge J. Llibre-Guerra, A. Matthew Prina, Adolfo Valhuerdi

**Author notes:** corresponding author Professor Martin Prince.

## Abstract

**Background:** The World Health Organization has reframed health and healthcare for older people around achieving the goal of healthy ageing. Recent evidence-based guidelines on Integrated Care for Older People focus on maintaining intrinsic capacity, addressing declines in mobility, nutrition, vision and hearing, cognition, mood and continence aiming to prevent or delay the onset of care dependence. The target group (with one or more declines in intrinsic capacity) is broad, and implementation at scale may be challenging in less-resourced settings.

Planning can be informed by assessing the prevalence of intrinsic capacity, characterising the target group, and validating the general approach by evaluating risk prediction for incident dependence and mortality.

**Methods:** Population-based cohort studies in urban sites in Cuba, Dominican Republic, Puerto Rico, Venezuela, and rural and urban sites in Peru, Mexico, India and China. Sociodemographic, behaviour and lifestyle, health, healthcare utilisation and cost questionnaires, and physical assessments were administered to all participants, with ascertainment of incident dependence, and mortality, three to five years later.

**Results:** In the 12 sites in eight countries, 17,031 participants were surveyed at baseline. Intrinsic capacity was least likely to be retained for locomotion (71.2%), vision (71.3%), cognition (73.5%), and mood (74.1%). Only 30% retained full capacity across all domains, varying between one quarter and two-fifths in most sites. The proportion retaining capacity fell sharply with increasing age, and declines affecting multiple domains were more common. Poverty, morbidity (particularly dementia, depression and stroke), and disability were concentrated among those with DIC, although only 10% were frail, and a further 9% had needs for care. Hypertension and lifestyle risk factors for chronic disease, healthcare utilization and costs were more evenly distributed in the general older population. 15,901 participants were included in the mortality cohort (2,602 deaths/ 53,911 person years of follow-up), and 12,965 participants in the dependence cohort (1900 incident cases/ 38,377 person-years). DIC (any decline, and number of domains affected) strongly and independently predicted incident dependence and death. Relative risks were higher for those who were frail, but were also substantially elevated for the much larger sub-groups yet to become frail. Mortality was mainly concentrated in the frail and dependent sub-groups.

**Conclusions:** Our findings support the strategy to optimize intrinsic capacity in pursuit of healthy ageing. Most needs for care arise in those with declines in intrinsic capacity who are yet to become frail. Implementation at scale requires community-based screening and assessment, and a stepped-care approach to intervention. Community healthcare workers’ roles would need redefinition to engage, train and support them in these tasks. ICOPE could be usefully integrated into community programmes orientated to the detection and case management of chronic diseases including hypertension and diabetes.

## Introduction

In May 2018, the World Health Organization (WHO) announced a new set of priorities - three ‘triple billion’ targets to ensure that by 2023 one billion more people benefit from universal health coverage, one billion more people have better protection from health emergencies, and one billion more people enjoy better health and well-being. Older people, who by 2019 will number one billion, typically experience worse health and health-related quality of life than younger persons yet do not access healthcare services proportionately, they are particularly vulnerable in emergency situations and account for half or more of all deaths after natural disasters (1). These disadvantages are most pronounced in low and middle income countries. Older peoples’ needs will therefore need to be accorded more priority. The WHO has also been active on this front, launching a landmark 2015 World Report on Ageing and Health (1), a Global Strategy and Action Plan on Ageing and Health (2016-2020)(2), and, in 2017, evidence-based guidelines on Integrated Care for Older People (ICOPE)(3).

The WHO World Report reframes health and healthcare for older people around the goal of healthy ageing, achieved when functional abilities, the “health related attributes that enable people to be and do what they have reason to value” are developed and maintained across the life course (1). At the core of functional ability are the physical, mental and cognitive intrinsic capacities of the individual, interacting with their environment. The ICOPE guidelines support the WHO Global Strategy core objective of ‘aligning health systems to the needs of older populations’. Implementation will promote community-level attention to declines in intrinsic capacity among older adults. Thirteen recommendations cover mobility loss, malnutrition, visual impairment and hearing loss, cognitive impairment and depressive symptoms, with additional modules on geriatric syndromes (urinary incontinence and risk of falls), and support for carers(3). WHO envisages brief screening for intrinsic capacity. Significant declines would prompt more comprehensive needs assessment. The emphasis is upon bringing assessment, and health and social care to the older person; in primary care, at home and in the community. Of necessity this involves task-shifting and task-sharing, particularly in low-resource settings where specialist older care services are underdeveloped. Given the extent of unmet needs, the optimal targeting of the ICOPE intervention will be an important consideration if health gains are to be realised in a cost-effective manner.

Aside from the evidence-base that supports each of the 13 ICOPE recommendations, the broad approach of multidimensional assessment and management is strongly supported, mainly by evidence from high income countries (HIC). The disability cascade is neither inevitable, unidirectional nor irreversible. Longitudinal studies of frailty in older populations show an overall tendency for stability and modest net progression, with little reversion from frail to completely robust states; however, significant numbers do improve, transiting from pre-frail to robust, and from frail to pre-frail states (4–9). Findings are similar from studies of transitions into and out of states of dependence (10–13), with some suggestion of less stability and more reversion in LMIC (11,12). More favourable trajectories are associated with more education (9,10,14), higher socioeconomic status (14), and adherence to healthy behaviours and lifestyles, particularly physical activity (13,15). Certain chronic progressive diseases (diabetes, cancer, COPD, osteoarthritis, dementia and cerebrovascular disease) are associated with a worse prognosis (5,16), and may mediate much of the effect of education (16). The critical time window for interventions, if any, has not yet been clearly established. A systematic review in 2008 identified 89 trials of home-based multi-dimensional assessment and intervention, all conducted in high income countries (HIC) (17). While, overall, there was evidence for improvements in physical function, fewer falls, and fewer admissions to nursing-homes and hospital, these benefits were mainly confined to interventions in the general older adult population, rather than amongst those selected because they were frail. This may have contributed to a widely held perception that early intervention is better, even necessary to prevent the onset of frailty and disability. However, more recent trials conducted in the Netherlands(18), Australia(19) and the USA(20) also indicate potential for improvements in health, functioning and quality of life among frail and dependent older people, with some evidence to support cost-effectiveness of multi-dimensional interventions (19,21).

To inform planning of ICOPE implementation activities we conducted a secondary analysis of data collected from the 10/66 Dementia Research Group population-based baseline and incidence wave surveys in 12 catchment areas in eight countries in Latin America, China and India. Our objectives were to:

1. determine the prevalence of intrinsic capacity overall, and in its several domains, clarifying the proportion of individuals likely to be identified with significant declines, and hence to be the target population for the ICOPE intervention
2. characterise the target group, and compare them with others, with for burden of disease (behavioural and lifestyle risk factors, morbidities and disability) and use of healthcare services
3. establish the extent of the overlap of decline in intrinsic capacity (DIC) with frailty and dependence (needs for care), and the concentration of burden indicators in each of these three groups when defined hierarchically
4. clarify the prospective association of DIC with incident dependence and mortality.

## Methods

### Settings and study design

The 10/66 Dementia Research Group’s (10/66 DRG) population-based studies of ageing and dementia in LMIC comprised baseline surveys of all older people aged 65 years and over living in geographically defined catchment areas in eight countries, with a follow-up three to five years later. For the current analysis this comprises urban and rural sites in Peru (Lima and Canete), Mexico (Mexico City and Morelos state), China (Xicheng and Daxing) and India (Chennai and Vellore), and urban sites in Cuba (Havana and Matanzas), Dominican Republic (Santo Domingo), Puerto Rico (Bayamon) and Venezuela (Caracas). Baseline population-based surveys were carried out between 2003 and 2007 and incidence wave follow-up assessments between 2008 and 2010. For India, the follow-up comprised a mortality sweep only, in the urban site. The protocols for the one phase surveys, comprised; a clinical interview; a health, medical history, healthcare utilisation and lifestyle interview; a cognitive assessment; a physical examination; and an informant interview, as detailed elsewhere(22,23). The study protocol and the consent procedures were approved by the King’s College London research ethics committee and in all countries where the research was carried out: 1-Medical Ethics Committee of Peking University the Sixth Hospital (Institute of Mental Health, China); 2- the Memory Institute and Related Disorders (IMEDER) Ethics Committee (Peru); 3-Finlay Albarran Medical Faculty of Havana Medical University Ethical Committee (Cuba); 4-Hospital Universitario de Caracas Ethics Committee (Venezuela); 5-Consejo Nacional de Bioética y Salud (CONABIOS, Dominican Republic); 6-Instituto Nacional de Neurología y Neurocirugía Ethics Committee (Mexico); 7-University of Puerto Rico, Medical Sciences Campus Institutional Review Board (IRB); 8 – Research and Ethics Committee, Christian Medical College (Vellore, India) and; 9. Institutional Ethics Committee, The Voluntary Health Services Multi Speciality Hospital and Research Centre (Chennai, India). Informed consent was documented in writing in all cases. Literate participants signed their consent. For participants who were illiterate, the information sheet was read to them in the presence of a literate independent witness, who attested by signature that this process had been completed, and that the participant had provided informed consent. For participants who lacked capacity to consent, agreement for their participation was obtained from next-of-kin. These procedures were approved by the ethics committees.

### Measures

Full details are available elsewhere(22). Here we summarise the measures directly relevant to the analyses presented in this paper.

Age was ascertained from participant and informant reports, and documented age, or an event calendar. Education level was self-reported, and coded as; no education, did not complete primary, completed primary, secondary, or tertiary education. Food insecurity was defined as reporting going hungry in the last one month, due to inadequate resources to procure food.

#### Intrinsic Capacities

Seven intrinsic capacities were evaluated, linked to the concepts and, where possible, the operationalisations current within the WHO ICOPE draft guidance on comprehensive assessment. 1. Neuromusculoskeletal Capacity - walking speed was assessed using a timed walking test (five metres at usual speed, turn, and return to the starting point); those completing in less than 16 seconds were considered to have capacity (allowing three seconds to make the turn, this corresponds to a walking speed of >0.8m/s). 2. Vitality - nutrition was considered adequate if the older person did not report weight loss of >=4.5kg in the last 3 months, and if their mid-upper arm circumference was measured to be >= 22 cms; this cut-point is used in the Mini Nutritional Assessment (MNA) ® to identify the most severe level of undernutrition(24). 3. Sensory Capacity (visual impairment) - vision was considered adequate when the older person did not report ‘eyesight problems’ that interfered with their activities to at least some extent, and they were not identified by the interviewer as being functionally blind. 4. Sensory Capacity (hearing loss) -hearing was considered adequate when the older person did not report ‘hearing problems or deafness’ that interfered with their activities to at least some extent, and they were not identified by the interviewer as being profoundly deaf. 5. Cognitive Capacity - cognitive function was assessed using the Community Screening Instrument for Dementia (CSI-D) COGSCORE, which tests multiple domains of cognitive function, and has been found to have robust cross-cultural measurement properties in the 10/66 study sites; those who scored >= 29.5 were considered to have cognitive capacity with scores below that threshold identifying ‘probable dementia’(25). 6. Psychological Capacity - psychological capacity was considered to be present if participants endorsed three or fewer of the 12 depression symptoms covered in the EURO-D depression scale; in previous analyses this cutpoint identified individuals with significant impairment of health related quality of life(26), although a higher cutpoint is better for identifying clinical cases(27). 7. Continence. Incontinence (either urinary, faecal or both) was established from informant report, and capacity was maintained if none of these were reported. Continence is not currently a primary focus of the ICOPE comprehensive assessment tool, although guidelines for the assessment and management of incontinence have been prepared by the Guideline Development Group.

#### Frailty

The physical frailty phenotype proposes five frailty indicators (exhaustion, weight loss, weak grip strength, slow walking speed and low energy expenditure). Individuals are frail if they meet three or more of the five criteria, pre-frail if they meet one or two, and non-frail if they meet none of the five criteria(28). We assessed four of the five indicators of frailty, but using a slightly different operationalisation to those originally proposed for exhaustion, weight loss and energy consumption, and omitting hand grip strength(29). As handgrip strength was not measured we considered participants frail if they fulfilled criteria for two or more of the four frailty indicators, and pre-frail if they met one; the effect is the same as imputing a value of one for handgrip strength. For most analyses we allocate frail or pre-frail individuals who already need care to the dependent sub-group, consistent with the concept of frailty as a vulnerability for disability and dependence.

#### Behavioural and lifestyle risk factors

Lifetime smoking history, current physical activity, alcohol consumption and dietary intake of fruit and vegetables were ascertained from self-report. Adverse behaviours were defined as current smoking, self-report of being ‘not very’ or ‘not at all’ physically active, weekly consumption of >= 28 units of alcohol for men and >=21 units for women, and fewer than four portions of fruit or vegetable in the last three days. Waist circumference was measured in centimetres using a flexible tape measure; central obesity was defined according to the criteria for metabolic syndrome in the Third Report of the National Cholesterol Education Program - a waist circumference of more than 101.6 centimetres in men and of more than 88.9 centimetres in women.

#### Morbidity

We assessed physical, mental and cognitive morbidity through measures of hypertension and diabetes; and the main contributors to disability and dependence(30,31); stroke, dementia and depression. Dementia was diagnosed according to the cross-culturally developed, calibrated and validated 10/66 dementia diagnosis algorithm(25). ICD-10 depressive episode was diagnosed using a computerised algorithm applied to the GMS structured clinical interview(32). Stroke was self-reported, but confirmed by the interviewer as having characteristic symptoms lasting for more than 24 hours(33). Hypertension was ascertained through blood pressure measurement applying WHO/ International Society of Hypertension criteria (SBP >=140 mm Hg and/ or diastolic blood pressure >= 90mm Hg) and self-report of previous diagnosis and treatment; those with elevated blood pressure were considered to be uncontrolled, regardless of detection or treatment(34).

#### Disability and dependence

Disability was assessed using the WHODAS 2.0 scale, developed by the WHO as a culture-fair assessment tool for use in cross-cultural comparative epidemiological and health services research(35,36). Dependence (needs for care) was interviewer coded after a series of open-ended probing questions to a key informant, and a detailed assessment of caregiving roles(31). To identify probable cases of incident dependence among participants who had died during the follow-up period, a predictive model for incident dependence was developed using variables from the informant section of the Community Screening Interview for Dementia (CSI-D) informant interview, which was available for all participants. For deceased participants this was conducted as part of an informant verbal autopsy interview, and referred to the period before death. The model used age, the total CSID informant score and the following items from the CSI-D informant interview: activity, feeding, toileting, dressing and household chores. The predictive model was developed from those who had survived, then applied to those who were deceased at follow-up to predict incident dependence.

#### Healthcare utilisation and costs

Details of healthcare cost estimations are provided elsewhere^18^. Participants were asked about contacts with primary healthcare professionals, public hospital doctors, other publically provided professionals, and private healthcare services (private doctors, dentists, and traditional healers). For each service, participants were asked how often they had used it in the last three months, the duration of the consultation, and fees for the service. Travel costs were also elicited. Lengths of stay and out of pocket costs for hospital admissions, and total costs of medication paid out-of-pocket for any of these services were also recorded. Out-of-pocket costs comprised the total annualised payments made by healthcare service users. Total costs from a public perspective reflect the actual cost to the provider, regardless of financing, including staff salaries, facilities and equipment, and overheads. Out-of-pocket costs and total costs were dichotomised at the 90^th^ centile of the distribution in each site to reflect catastrophic healthcare spending and high total healthcare costs respectively.

### Analysis

We report, descriptively, the prevalence of retained intrinsic capacity for each of the seven domains by site, and, for the whole sample, prevalence by five-year age group from 65-69 years to 90 years and over. We describe the sociodemographic and health characteristics (lifestyle risk factors, morbidity, disability and needs for care), healthcare utilisation and costs of those with declines in intrinsic capacity (DIC) affecting one or more domain, further stratified by frailty status and needs for care. Characteristics are compared with those with fully retained capacity (chi squared tests and t-tests), and across all four strata (full capacity/ DIC only/ DIC and pre-frail/ DIC and frail/ DIC and dependent - chi squared and oneway ANOVA tests for trend).

We modelled the effect of intrinsic capacity exposures on the incidence of dependence (or ‘probable dependence’ among those who had died) using a competing-risks regression derived from Fine and Gray’s proportional subhazards model [27] (Stata stcrreg command), based on a cumulative incidence function, indicating the probability of failure (onset of dependence) before a given time, acknowledging the possibility of a competing event (dependence-free death), and reporting adjusted sub-hazard ratios (aSHR). Competing risks regression keeps those who experience competing events at risk so that they can be counted as having no chance of failing. We modelled the effect of intrinsic capacity on mortality using Cox’s Proportional Hazards, generating cumulative survival probability curves and reporting adjusted hazard ratios (aHR). Proportional hazards assumptions were checked using methods based on Schoenfeld residuals (Stata phtest command), and fitted interactions with time for covariates that violated these assumptions. Time to death was the time from baseline interview to the exact date of death. Time to dependence onset (which could not be ascertained precisely) was the midpoint between baseline and follow-up interview, or death. All effect sizes are presented with robust 95% confidence intervals adjusted for household clustering.

Models comprised 1. The effect of any DIC, 2. The effect of DIC when stratified as; DIC only/ DIC and pre-frail/ DIC and frail/ and (mortality model only) DIC and dependent, 3. The effect of DIC per number of intrinsic capacity domains affected, before and after controlling for frailty (and dependence in the mortality model). All models were adjusted for age, sex, and education, and estimated separately for each site with the results combined using a fixed effects meta-analysis. Higgins I^2^ estimates the proportion of between-site variability in the estimates accounted for by heterogeneity, as opposed to sampling error; up to 40% heterogeneity is conventionally considered negligible, while up to 60% reflects moderate heterogeneity [28].

## Results

### Sample characteristics

In all, 17,031 participants were surveyed at baseline in the 12 sites in eight countries. Their characteristics have been reported in detail elsewhere (23). Mean ages varied between 71.3 and 76.3 years, higher in urban than rural and in more than less developed sites (supplementary Table 1). Most participants (62.4%) were female. Education levels varied widely among sites, with between 14.4% and 90.7% having completed primary education, lowest in rural sites in India, Mexico and China and in the Dominican Republic, and highest in urban Peru, Puerto Rico and Cuba. Food insecurity was commonest in urban (20.8%) and rural India (14.1%), in rural Peru (13.5%) and Dominican Republic (12.1%). Overall, 16.1% reported three or more physical impairments, 6.7% had a history of stroke, 5.5% met criteria for ICD-10 Depressive episode in the last one month, and 9.3% for 10/66 Dementia diagnosis. Physical impairments and stroke were less frequently reported in rural and less developed sites, and depression was rarely identified in China.

**Table 1.** Prevalence of individual capacities, and full capacity, by site

|  | Neuromusculoskeletal capacity (%) | Vitality (nutrition) | Sensory capacity (vision) | Sensory capacity (hearing) | Cognitive capacity | Psychological capacity | Continence | Full capacity |
| --- | --- | --- | --- | --- | --- | --- | --- | --- |
| Cuba | 1713 (58.6%) | 2580 (87.8%) | 2063 (70.4%) | 2626 (89.5%) | 2287 (77.7%) | 2210 (76.4%) | 2801 (96.1%) | 842 (28.6%) |
| Dominican Republic | 807 (42.1%) | 1671 (83.7%) | 1196 (59.6%) | 1659 (82.6%) | 1404 (69.8%) | 1237 (62.0%) | 1912 (95.2%) | 301 (15.0%) |
| Puerto Rico | 1292 (85.0%) | 1479 (90.0%) | 1467 (73.3%) | 1645 (82.2%) | 1495 (74.4%) | 1578 (82.7%) | 1913 (95.7%) | 791 (39.4%) |
| Peru (urban) | 894 (65.1%) | 1116 (81.4%) | 921 (66.9%) | 1070 (77.8%) | 1167 (84.5%) | 945 (70.8%) | 1317 (95.5%) | 384 (27.8%) |
| Peru (rural) | 411 (74.7%) | 444 (80.6%) | 352 (63.9%) | 457 (82.9%) | 436 (79.0%) | 404 (73.7%) | 543 (98.7%) | 143 (25.9%) |
| Venezuela | 1188 (81.5%) | 1304 (81.5%) | 1155 (59.8%) | 1632 (84.3%) | 1617 (82.3%) | 1371 (70.5%) | 1900 (96.8%) | 609 (31.0%) |
| Mexico (urban) | 883 (93.9%) | 888 (88.9%) | 716 (71.4%) | 799 (79.7%) | 728 (72.6%) | 684 (68.7%) | 975 (97.2%) | 342 (34.1%) |
| Mexico (rural) | 854 (93.5%) | 806 (80.8%) | 641 (64.2%) | 769 (76.9%) | 617 (61.7%) | 731 (73.8%) | 982 (98.2%) | 252 (25.2%) |
| China (urban) | 1021 (88.0%) | 1136 (98.4%) | 962 (82.9%) | 1013 (81.3%) | 1048 (90.3%) | 1082 (96.1%) | 1099 (94.9%) | 728 (62.8%) |
| China (rural) | 338 (33.8%) | 989 (98.7%) | 937 (93.5%) | 911 (90.9%) | 820 (81.8%) | 959 (98.5%) | 981 (97.9%) | 262 (26.1%) |
| India (urban) | 906 (90.9%) | 657 (66.0%) | 913 (90.9%) | 971 (96.8%) | 574 (57.1%) | 610 (61.0%) | 975 (98.1%) | 275 (27.4%) |
| India (rural) | 907 (90.8%) | 647 (66.1%) | 776 (77.7%) | 844 (84.5%) | 332 (33.2%) | 532 (55.8%) | 978 (98.1%) | 120 (12.0%) |
| Total | 11214 (71.2%) | 13717 (84.5%) | 12099 (71.3%) | 14396 (84.8%) | 12525 (73.5%) | 12343 (74.1%) | 16376 (96.5%) | 5049 (29.6%) |

### Prevalence of intrinsic capacity

In the pooled data set, capacity was least likely to be retained with respect to locomotion (71.2%), vision (71.3%), cognition (73.5%), and mood (74.1%), and most likely to be retained for continence (96.5%). Just 29.6% retained full capacity across all domains (Table 1). This proportion varied between one quarter and two-fifths in most sites with low outliers in rural India (11.0%) and Dominican Republic (15.0%) and a high outlier in urban China (62.8%). There was considerable variation in the prevalence of individual capacities among sites, mostly arising from the China sites; walking speeds were exceptionally low in rural China, cognition well preserved in urban China, and undernutrition and low mood were rarely encountered in either site.

The proportion retaining capacity decreased significantly with age for all individual capacities studied, other than psychological capacity (Fig 1). For continence, hearing and vision, declines in capacity were more pronounced after the age of 80 years. The proportion retaining full capacity declined linearly with age from 38.9% at ages 65-69 years, to 3.6% for those aged 90 years and over. Loss of capacity in multiple domains was also more common in older age groups (Fig 2); loss of capacity in four or more domains was detected in 11.3% of those aged 65-69 years, and in 60.2% of those aged 90 years or over.

**Fig 1.**
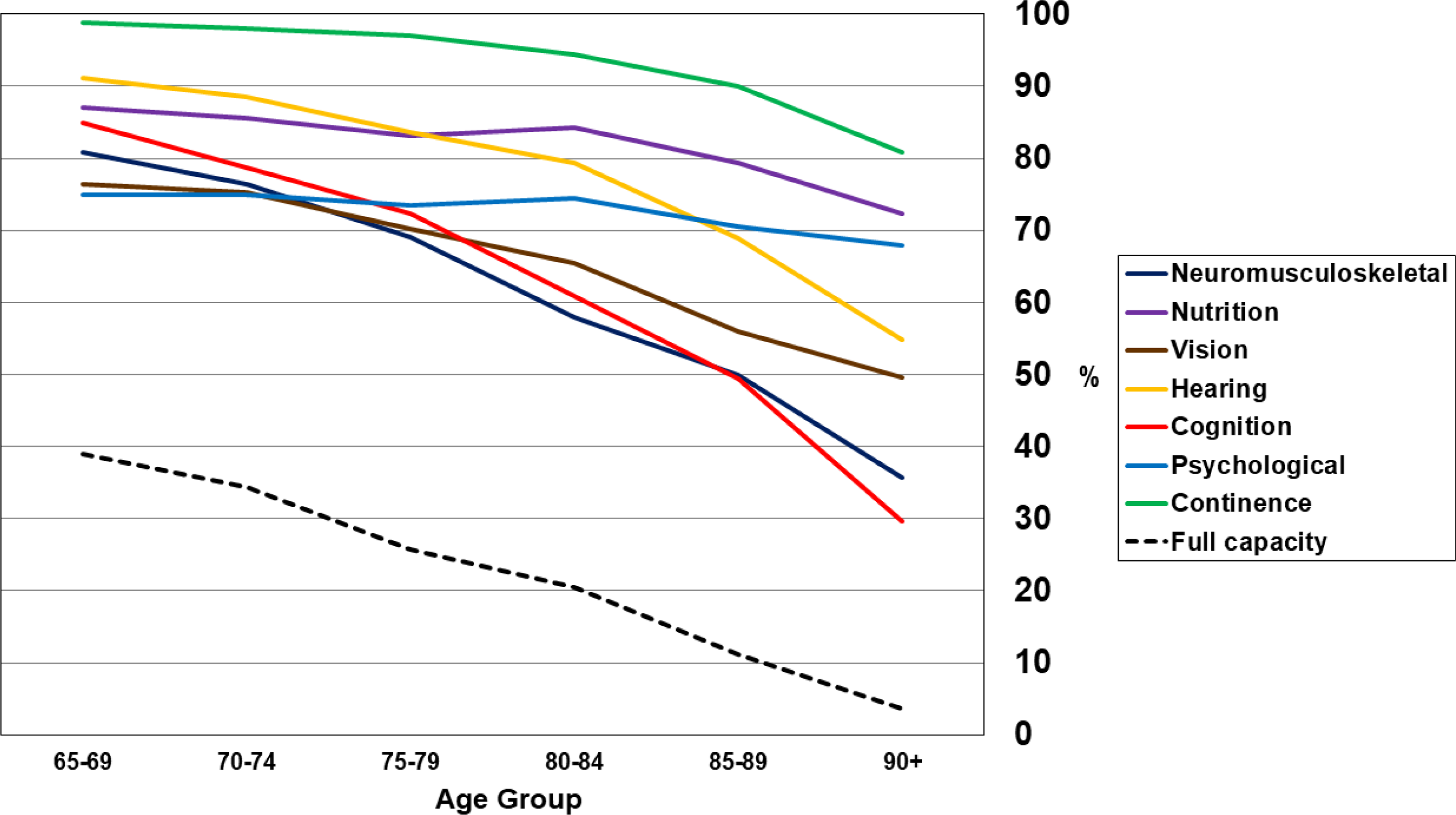
Prevalence (%) of retained intrinsic capacity, by age group

**Fig 2.**
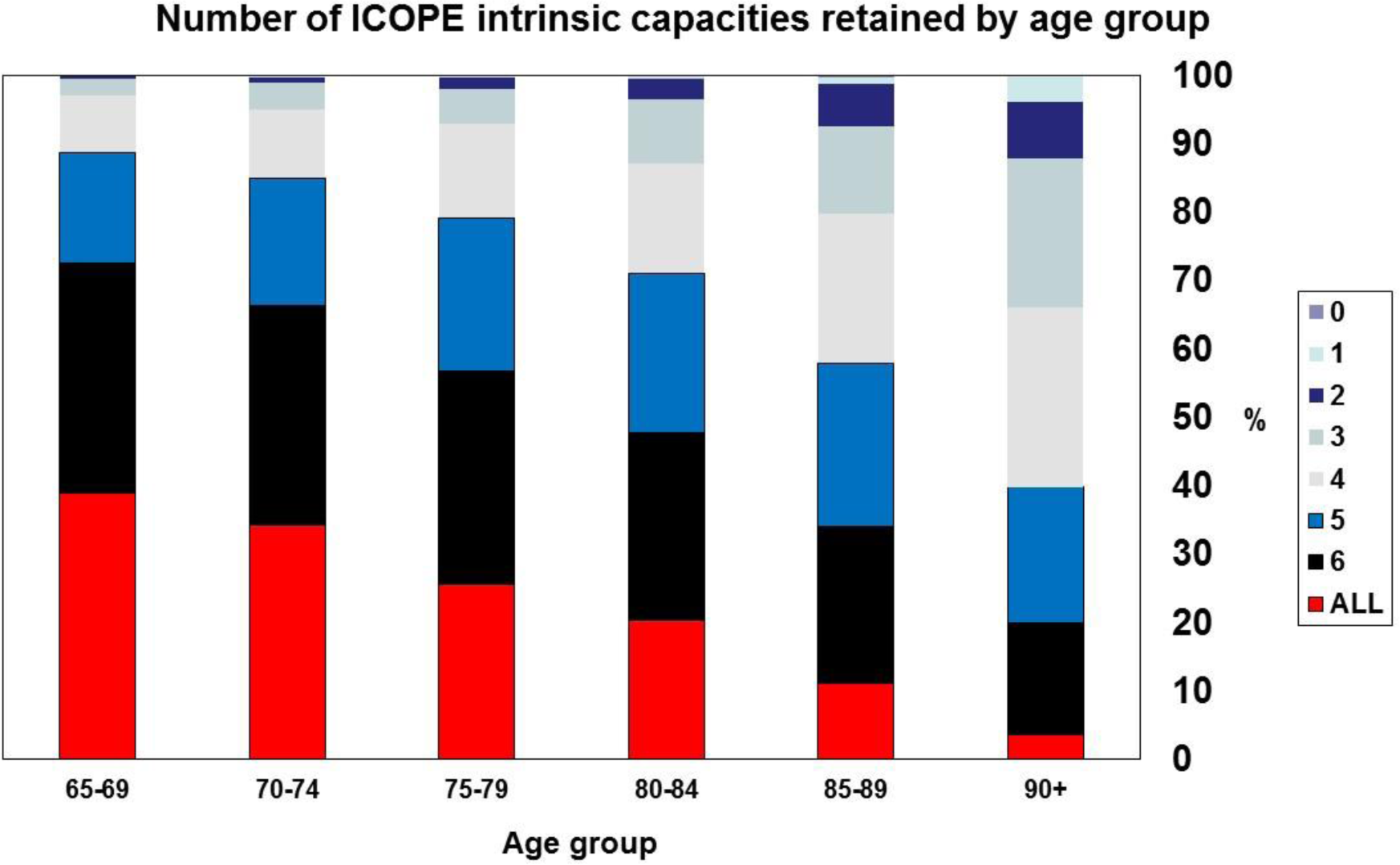
Distribution (%) of number of intrinsic capacities retained, by age group

### Characteristics of those with declines in intrinsic capacity

In principle, all those with declines in intrinsic capacity (DIC) affecting one or more domain might benefit from the ICOPE intervention (the target group). Overall, 11,982 older people (70.4%) met this criterion, the proportion varying between 37.2% (urban China) and 88.0% (rural India) by site, with other sites clustering closely around the median of 72.4% (25^th^ centile 67.4%, 75^th^ 74.4%). Among those with DIC, 4,541 (37.9%) were neither frail nor in need of care, a further 4,193 (35.0%) were pre-frail, 1743 (10.2%) were frail, and 1,505 (8.8%) had established needs for care. The characteristics of the target group are shown in Table 2, further broken down by frailty status and needs for care. Those targeted were on average three years older than those not selected, and mean age increased monotonically from the DIC only, to the pre-frail, frail and dependent groups. Women were over-represented. Poverty and disadvantage were particularly marked; those in the DIC target group had nearly three times the prevalence of food insecurity, and most had not completed primary education. Morbidity and disability are also concentrated among those in the target group; this was very marked for depression, dementia, and stroke, and these conditions were further concentrated among those who were also frail or had needs for care. Diabetes, health service utilisation and costs were moderately associated with DIC. Other than physical activity and diet, lifestyle risk factors for chronic disease were not concentrated among those with DIC.

**Table 2.** Characteristics of the target group with declines in intrinsic capacity (DIC), stratified by frailty states and dependence

|  | No DIC<br>N=5049 | DIC<br>(target<br>group)<br>n=11982 | DIC, with<br>no frailty<br>or needs<br>for care<br>n=4541 | DIC and<br>pre-frail<br>n=4193 | DIC and<br>frail<br>n=1743 | DIC and<br>needing<br>care<br>n=1505 | Chi<br>square<br>or F<br>statistic<br>(1df) | Chi<br>square<br>or F<br>statistic<br>(1df,<br>(TFT) |
| --- | --- | --- | --- | --- | --- | --- | --- | --- |
| Age in years | 72.0 (5.6) | 75.1<br>(7.3) | 73.7 (6.5) | 74.3<br>(7.0) | 75.9<br>(7.2) | 80.1<br>(8.0) | 691.7<br>p<0.001 | 464.2<br>p<0.001 |
| Male gender | 2285<br>(45.4%) | 4063<br>(34.3%) | 1654<br>(37.7%) | 1366<br>(32.6%) | 568<br>(32.6%) | 475<br>(31.6%) | 183.3<br>p<0.001 | 176.8<br>p<0.001 |
| Did not<br>complete<br>primary | 1495<br>(29.8%) | 5997<br>(50.9%) | 2274<br>(51.9%) | 2060<br>(49.2%) | 866<br>(49.9%) | 797<br>(53.7%) | 636.4<br>p<0.001 | 358.1<br>p<0.001 |
| Food<br>insecurity | 146<br>(2.9%) | 991<br>(8.5%) | 235<br>(5.4%) | 400<br>(9.6%) | 231<br>(13.4%) | 125<br>(8.4%) | 168.6<br>p<0.001 | 223.4<br>p<0.001 |
| ICD 10<br>Depression | 3 (0.1%) | 934<br>(7.9%) | 96 (2.2%) | 324<br>(7.7%) | 305<br>(17.5) | 209<br>(13.9%) | 413.3<br>p<0.001 | 961.3<br>p<0.001 |
| Dementia | 21 (0.4%) | 1483<br>(12.5%) | 252<br>(5.7%) | 293<br>(7.0%) | 190<br>(10.9%) | 748<br>(49.8%) | 638.0<br>p<0.001 | 2350.2<br>p<0.001 |
| Stroke | 172<br>(3.4%) | 929<br>(7.9%) | 188<br>(4.3%) | 259<br>(6.2%) | 146<br>(8.4%) | 336<br>(22.5%) | 113.2<br>p<0.001 | 521.7<br>p<0.001 |
| COPD | 148<br>(2.9%) | 661<br>(5.6%) | 176<br>(4.0%) | 244<br>(5.8%) | 134<br>(7.7%) | 107<br>(7.2%) | 53.7<br>p<0.001 | 96.8<br>p<0.001 |
| Diabetes | 743<br>(14.8%) | 1999<br>(16.9%) | 550<br>(12.5%) | 752<br>(18.0%) | 356<br>(20.5%) | 341<br>(22.9%) | 11.9<br>p<0.001 | 94.2<br>p<0.001 |
| Hypertension | 3262<br>(66.2%) | 7944<br>(68.4%) | 2887<br>(66.9%) | 2800<br>(67.9%) | 1224<br>(70.9%) | 1033<br>(70.9%) | 7.0<br>p=0.008 | 19.1<br>p<0.001 |
| Uncontrolled<br>hypertension | 2025<br>(41.1%) | 4928<br>(43.7%) | 1969<br>(46.7%) | 1646<br>(41.3%) | 714<br>(42.5%) | 599<br>(42.8%) | 1.5<br>p=0.21 | 1.7<br>p=0.19 |
| Obesity | 1970<br>(41.5%) | 4304<br>(38.5%) | 1434<br>(34.0%) | 1632<br>(41.1%) | 705<br>(42.3%) | 533<br>(39.7%) | 12.9<br>p<0.001 | 1.1<br>p=0.30 |
| Low physical<br>activity | 889<br>(17.7%) | 4795<br>(40.7%) | 1183<br>(27.0) | 1515<br>(36.3) | 926<br>(53.3%) | 1171<br>(78.6%) | 831.0<br>p<0.001 | 2167.3<br>p<0.001 |
| Less than<br>daily fruit<br>and veg | 891<br>(17.8%) | 2974<br>(25.4%) | 893<br>(20.5%) | 1172<br>(28.2%) | 516<br>(30.0%) | 393<br>(26.6%) | 112.6<br>p<0.001 | 160.5<br>p<0.001 |
| Hazardous<br>alcohol use | 131<br>(2.8%) | 344<br>(3.1%) | 142<br>(3.4%) | 109<br>(2.8%) | 53<br>(3.3%) | 40<br>(2.8%) | 0.96<br>p=0.33 | 0.02<br>p=0.88 |
| Current<br>smoker | 738<br>(14.7%) | 1801<br>(15.3%) | 823<br>(18.8%) | 569<br>(13.6%) | 252<br>(14.5%) | 157<br>(10.5%) | 6.3<br>p=0.04 | 86.2<br>p<0.001 |
| Used health<br>care services<br>in last 3<br>months | 2348<br>(46.6%) | 6573<br>(55.6%) | 2176<br>(49.6%) | 2437<br>(58.1%) | 1084<br>(62.2%) | 876<br>(58.2%) | 112.2<br>p<0.001 | 181.1<br>p<0.001 |
| Out-of-pocket healthcare cost (top tenth by site) | 414<br>(8.2%) | 1295<br>(11.0%) | 350<br>(8.0%) | 446<br>(10.6%) | 222<br>(12.7%) | 277<br>(18.5%) | 28.4<br>p<0.001 | 140.2<br>p<0.001 |
| Total healthcare costs (top tenth by site) | 281<br>(6.6%) | 1172<br>(11.0%) | 338<br>(8.4%) | 407<br>(10.8%) | 202<br>(13.0%) | 225<br>(17.3%) | 65.6<br>p<0.001 | 158.3<br>p<0.001 |
| Pain scale – mean (SD) | 2.7 (3.1) | 4.2 (4.0) | 3.3 (3.4) | 4.5 (4.0) | 5.5 (4.2) | 4.1 (4.5) | 537.5<br>p<0.001 | 261.6<br>p<0.001 |
| WHODAS 2.0 disability – mean (SD) | 3.8 (7.9) | 17.0<br>(20.8) | 8.5 (12.0) | 13.0<br>(14.6) | 21.3<br>(18.9) | 48.0<br>(13.0) | 1802.3<br>p<0.001 | 2848.1<br>p<0.001 |

### Predictors of incident dependence

12,965 participants who had no needs for care at baseline were included in the baseline survey, of whom 9,066 (69.9%) were re-interviewed, and 1,686 (13.0%) had died. Enough information was available to allocate an outcome of dependence-free death (competing risk) to 832 individuals. Onset of dependence before death was defined post mortem in 498, and a further 1402 incident cases were identified among those re-interviewed. Therefore, 1900 incident cases were identified in 38,377 person years of follow-up, a rate of 49.5 per 1000 person years.

Mutually controlling for all other domains of intrinsic capacity, and further adjusting for age, gender and education, declines in cognitive capacity (pooled adjusted subhazard ratio [aSHR] 2.56, 95% CI 2.26-2.90) and continence (pooled aSHR 3.32, 95% CI 2.48-4.44) were strongly associated with incident dependence (Table 2). Decline in psychological capacity (pooled aSHR 1.33, 95% CI 1.18-1.51), locomotor capacity (pooled aSHR 1.42, 95% CI 1.26-1.60) and nutrition (pooled aSHR 1.21, 95% CI 1.04-1.41) were moderately associated. Neither of the two sensory capacities (vision and hearing) was associated with incident dependence.

**Table 2.** Adjusted^1^ associations of individual declines in intrinsic capacity with incident dependence

| Catchment area | Cognitive | Psychological | Continence | Locomotor | Hearing | Vision | Nutrition |
| --- | --- | --- | --- | --- | --- | --- | --- |
| Cuba | 2.93 (2.14-4.01) | 1.40 (1.03-1.88) | 1.81 (0.58-5.63) | 1.46 (1.11-1.91) | 0.99 (0.68-1.46) | 0.93 (0.69-1.23) | 1.39 (0.99-1.95) |
| Dominican Republic | 2.51 (1.95-3.23) | 1.14 (0.89-1.45) | 3.63 (1.86-7.10) | 1.25 (0.99-1.58) | 0.99 (0.76-1.29) | 1.07 (0.85-1.33) | 1.33 (1.00-1.76) |
| Puerto Rico | 3.23 (2.30-4.55) | 1.43 (0.98-2.08) | 1.93 (1.14-3.29) | 1.68 (1.20-2.36) | 0.96 (0.66-1.41) | 1.07 (0.77-1.24) | 0.93 (0.55-1.59) |
| Peru Urban | 4.03 (2.47-6.55) | 1.20 (0.80-1.81) | 9.07 (4.82-17.09) | 1.16 (0.79-1.72) | 0.88 (0.55-1.41) | 1.14 (0.76-1.71) | 1.00 (0.61-1.65) |
| Peru Rural | 3.93 (1.69-9.13) | 0.60 (0.27-1.32) | DNC | 1.33 (0.69-2.56) | 1.13 (0.50-2.56) | 1.55 (0.81-2.98) | 1.53 (0.72-3.24) |
| Venezuela | 2.52 (1.76-3.61) | 1.47 (1.02-2.10) | 2.93 (1.36-6.32) | 1.93 (1.37-2.72) | 1.32 (0.87-2.02) | 1.18 (0.85-1.63) | 1.16 (0.79-1.72) |
| Mexico Urban | 1.91 (1.15-3.16) | 1.43 (0.93-2.20) | 4.42 (1.39-14.05) | 1.61 (0.80-3.22) | 0.69 (0.40-1.20) | 1.34 (0.87-2.09) | 0.82 (0.37-1.82) |
| Mexico Rural | 2.33 (1.46-3.74) | 1.55 (0.98-2.47) | DNC | 1.89 (1.03-3.46) | 0.97 (0.59-1.58) | 1.00 (0.66-1.52) | 1.16 (0.69-1.96) |
| China Urban | 0.76 (0.35-1.65) | 1.71 (0.67-4.36) | 1.50 (0.31-7.35) | 1.65 (0.97-2.80) | 1.41 (0.95-2.09) | 1.28 (0.89-1.84) | DNC |
| China Rural | 1.98 (1.36-2.88) | 3.94 (1.70-9.13) | DNC | 1.05 (0.73-1.51) | 0.99 (0.58-1.71) | 1.20 (0.65-2.21) | DNC |
| **Pooled** | **2.56 (2.26-2.90)**<br>**54 (6-77)** | **1.33 (1.18-1.51)**<br>**33 (0-68)** | **3.32 (2.48-4.44)**<br>**63 (16-84)** | **1.42 (1.26-1.60)**<br>**12 (0-52)** | **1.03 (0.90-1.17)**<br>**0 (0-62)** | **1.11 (0.99-1.24)**<br>**0 (0-62)** | **1.21 (1.04-1.41)**<br>**0 (0-68)** |

1. Controlling for age, gender, education and all other capacity losses
DNC = Did not converge

DIC affecting one or more domain was associated with incident independence (pooled aSHR 1.91, 95% CI 1.69-2.17) (Table 3). Although the increased risk was somewhat concentrated among those who were frail (pooled aSHR 2.90, 95% CI 2.44-3.45), it was still apparent among the larger sub-groups with DIC who were neither pre-frail nor frail (pooled aSHR 1.72, 95% CI 1.50-2.17), and who were considered pre-frail (pooled aSHR 1.89, 95% CI 1.64-2.19).

**Table 3.** The associations of declines in intrinsic capacity (DIC), with incident dependence, stratified by frailty status

|  | No DIC<br>n=5049 | DIC n=11,982 | DIC, with no<br>frailty or needs<br>for care<br>N=4,541 | DIC and pre-frail<br>n=4,193 | DIC and frail<br>N=1,743 |
| --- | --- | --- | --- | --- | --- |
| Cuba | 1 (ref) | 2.71 (1.89-3.91) | 2.36 (1.59-3.50) | 2.75 (1.84-4.13) | 3.61 (2.35-5.56) |
| Dominican Republic | 1 (ref) | 1.51 (1.08-2.13) | 1.37 (0.95-1.98) | 1.49 (1.04-2.15) | 1.88 (1.26-2.78) |
| Puerto Rico | 1 (ref) | 2.21 (1.67-2.94) | 2.13 (1.50-3.02) | 2.08 (1.49-2.90) | 2.68 (1.83-3.94) |
| Peru Urban | 1 (ref) | 3.05 (1.42-6.52) | 2.83 (1.26-6.38) | 2.80 (1.25-6.29) | 3.68 (1.63-8.31) |
| Peru Rural | 1 (ref) | 2.91 (0.99-8.56) | 2.91 (0.91-9.29) | 1.91 (0.60-6.06) | 6.22 (1.90-20.44) |
| Venezuela | 1 (ref) | 2.27 (1.59-3.23) | 1.82 (1.21-2.74) | 2.25 (1.53-3.32) | 3.86 (2.46-6.04) |
| Mexico Urban | 1 (ref) | 1.89 (1.17-3.06) | 1.51 (0.82-2.80) | 1.60 (0.94-2.73) | 3.61 (1.98-6.59) |
| Mexico Rural | 1 (ref) | 1.84 (1.02-3.30) | 1.46 (0.76-2.81) | 1.55 (0.81-2.98) | 3.40 (1.77-6.56) |
| China Urban | 1 (ref) | 1.56 (1.17-2.09) | 1.54 (1.11-2.14) | 1.76 (1.17-2.64) | 1.12 (0.38-3.33) |
| China Rural | 1 (ref) | 1.30 (0.87-1.95) | 1.27 (0.84-1.93) | 1.26 (0.77-2.08) | 2.37 (1.83-3.94) |
| **Pooled** | **1 (ref)** | **1.91 (1.69-2.17)**<br>**39 (0-71)** | **1.72 (1.50-1.98)**<br>**15 (0-56)** | **1.89 (1.64-2.19)**<br>**12 (0-54)** | **2.90 (2.44-3.45)**<br>**32 (0-68)** |

When comparing the crude and mutually adjusted effects of frailty and number of DICs, number of DICs emerged as the more independent predictor of incident dependence. The effect of frailty was considerably attenuated after controlling for DIC (from pooled aSHR 1.78 to aSHR 1.21), whereas that of number of DICs was only moderately reduced when controlling for frailty (from pooled aSHR 1.35 per DIC to aSHR 1.32) (Table 4).

**Table 4.**
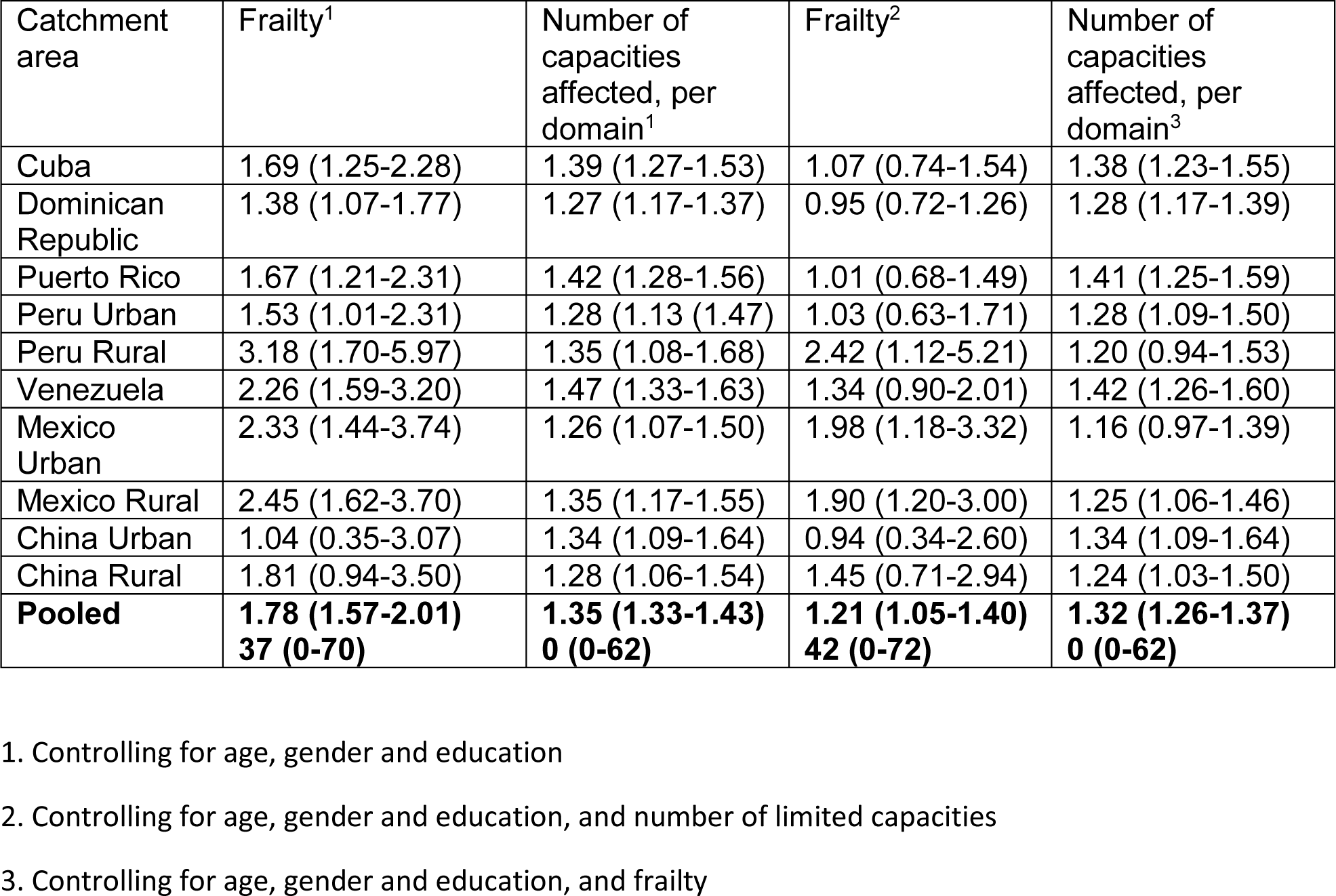
The associations of frailty and declines of intrinsic capacity (number of domains affected, per domain) with incident dependence

| Catchment area | Frailty<sup>1</sup> | Number of capacities affected, per domain<sup>1</sup> | Frailty<sup>2</sup> | Number of capacities affected, per domain<sup>3</sup> |
| --- | --- | --- | --- | --- |
| Cuba | 1.69 (1.25-2.28) | 1.39 (1.27-1.53) | 1.07 (0.74-1.54) | 1.38 (1.23-1.55) |
| Dominican Republic | 1.38 (1.07-1.77) | 1.27 (1.17-1.37) | 0.95 (0.72-1.26) | 1.28 (1.17-1.39) |
| Puerto Rico | 1.67 (1.21-2.31) | 1.42 (1.28-1.56) | 1.01 (0.68-1.49) | 1.41 (1.25-1.59) |
| Peru Urban | 1.53 (1.01-2.31) | 1.28 (1.13-1.47) | 1.03 (0.63-1.71) | 1.28 (1.09-1.50) |
| Peru Rural | 3.18 (1.70-5.97) | 1.35 (1.08-1.68) | 2.42 (1.12-5.21) | 1.20 (0.94-1.53) |
| Venezuela | 2.26 (1.59-3.20) | 1.47 (1.33-1.63) | 1.34 (0.90-2.01) | 1.42 (1.26-1.60) |
| Mexico Urban | 2.33 (1.44-3.74) | 1.26 (1.07-1.50) | 1.98 (1.18-3.32) | 1.16 (0.97-1.39) |
| Mexico Rural | 2.45 (1.62-3.70) | 1.35 (1.17-1.55) | 1.90 (1.20-3.00) | 1.25 (1.06-1.46) |
| China Urban | 1.04 (0.35-3.07) | 1.34 (1.09-1.64) | 0.94 (0.34-2.60) | 1.34 (1.09-1.64) |
| China Rural | 1.81 (0.94-3.50) | 1.28 (1.06-1.54) | 1.45 (0.71-2.94) | 1.24 (1.03-1.50) |
| **Pooled** | **1.78 (1.57-2.01)**<br>**37 (0-70)** | **1.35 (1.33-1.43)**<br>**0 (0-62)** | **1.21 (1.05-1.40)**<br>**42 (0-72)** | **1.32 (1.26-1.37)**<br>**0 (0-62)** |

1. Controlling for age, gender and education
2. Controlling for age, gender and education, and number of limited capacities
3. Controlling for age, gender and education, and frailty

### Predictors of mortality

15,901 participants were included in the mortality cohort, of whom vital status at follow-up was ascertained for 13,936 (87.5%). 2,602 deaths were recorded during 53,911 person years of follow-up, a mortality rate of 48.2 per 1,000 person years. Having controlled for age, gender and education, mortality was strongly influenced by DIC with negligible heterogeneity across sites (HR 1.66, 95% CI 1.49-1.85, I^2^=0, 0-60). Mortality risk was considerably elevated in the frail (HR 2.20, 95% CI 1.89-2.56, I^2^=0, 0-60) and dependent (HR 3.92, 95% CI 3.43-4.49, I^2^=33, 0-67) sub-groups with only around half surviving to six years (Fig 3).

**Figure 3.**
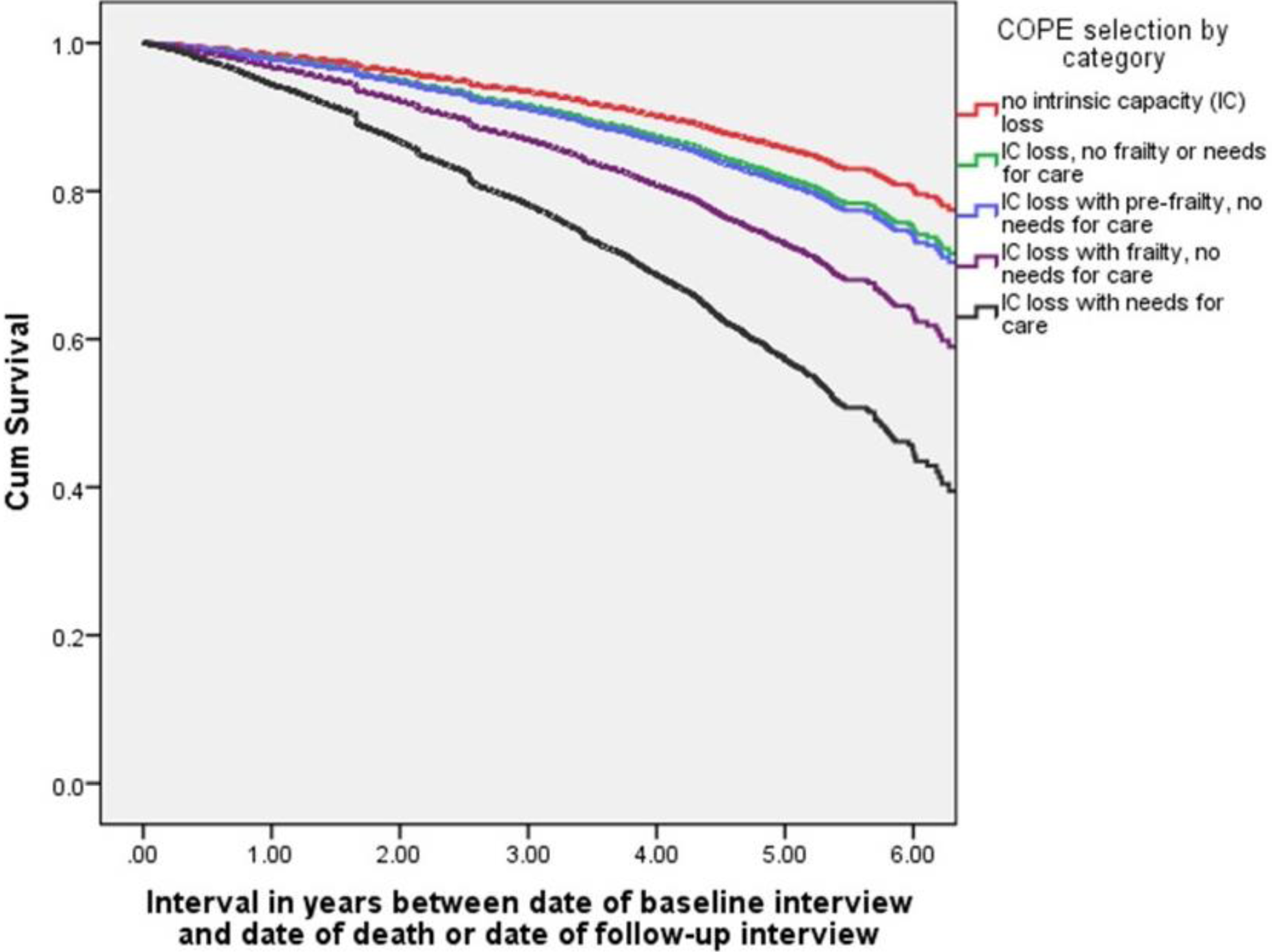
Cumulative survival probability by intrinsic capacity, frailty and needs for care

However, there was also a small increase in mortality in those with DIC who were neither pre-frail nor frail (HR 1.27, 95% CI 1.12-1.45, I^2^=39, 0-70); and in those who were pre-frail (HR 1.42, 95% CI 1.24-1.63, I^2^=1, 0-61). Although mortality risk increased monotonically with each additional DIC (HR 1.32, 95% CI 1.28-1.36, I^2^=44, 0-72), this effect was substantially attenuated after controlling for frailty and dependence (HR 1.16, 95% CI 1.12-1.20, I^2^=58, 17-78). The proportional hazards test suggested some departures from proportional hazard assumptions in some sites. These all related to control covariates (age, or age and gender), and not to the DIC variables. Interactions with time were introduced for those covariates for the sites where this was an issue, and the effects on the pooled estimates were negligible.

## Discussion

In this prospective population-based cohort study, we have shown that, typically, two-thirds to three-quarters of those aged 65 years and over have experienced declines affecting one or more domain of intrinsic capacity (DIC). DIC are more prevalent with increasing age, as are DIC affecting multiple domains. While DIC is strongly associated with disability, only around one fifth of those affected are frail or dependent. There is a strong concentration of cases of dementia, stroke and clinically relevant depression among those with DIC, particularly among those who are already frail or need care. Hypertension, diabetes, and behavioural risk factors for chronic disease are less strongly associated with intrinsic capacity, with a substantial prevalence among those who are yet to have experienced DIC. DIC (both any decline and number of domains affected) strongly and independently predict incident dependence and death. The increased risks of dependence are concentrated among those who are frail, but, nevertheless, are also substantially elevated for the much larger sub-groups who are yet to become frail. Mortality is particularly high in the frail and dependent groups.

### Strengths and weaknesses

The main strengths of this study include the large population-based samples with a high proportion of responders and modest missing data. The diverse countries and settings, across three continents, allowed us to study and explore variation in the context of a common study protocol, with detailed assessment of physical, mental and cognitive morbidities, healthcare utilisation, disability, frailty and needs for care. The WHO has yet to finalise the criteria that it will recommend for screening for intrinsic capacity. Both the approach, and the threshold selected are likely to have important impacts on the proportions that screen positive. Informant reports, of, for example, incontinence may only identify more severe cases; hence, perhaps, the low prevalence and the strong association with both prevalent and incident dependence in our study. Vision and hearing would be better assessed through objective tests of functioning, rather than the self-report and interviewer judgment that we had available. As previously reported our frailty indicators were operationalised in slightly different ways(29) from those originally suggested(28), and consequent measurement error may have compromised their ability to predict the onset of needs for care.

### Implications for policy and practice

At baseline, we identified those with declines in intrinsic capacity (DIC), and further subdivided these into four groups reflecting the common concept of the ‘disability cascade’ through which, with the accumulation and progression of DIC individuals become frail, experience limitation in core activities of daily living, and require care. The findings from our analysis broadly support the WHO’s strategy to focus upon optimising intrinsic capacity in pursuit of healthy ageing. If the ultimate public health aim is to reduce the future toll of disability and dependence, then most incident cases will arise among those with DIC who are yet to become frail, and a simple count of domains of intrinsic capacity affected is the better predictor of the onset of needs for care across the spectrum. Our research confirms the importance of cognitive and mental health in the maintenance of functional independence, and the inclusion of cognitive and psychological capacity in the intrinsic capacity framework ensures that attention is given to brain and mind, alongside physical frailty. Nevertheless, the aim to optimise intrinsic capacity broadens the potential scope of the intervention from the one-fifth of the older population who are already frail or dependent (the focus of most programs to date), to encompass up to three-quarters of the general population of older people. This raises important questions of appropriate targeting, from perspectives of feasibility and cost-effectiveness.

A comprehensive approach to healthy ageing should involve promotion (optimising health behaviours and structural conditions in society), prevention (managing underlying risk factors and conditions), treatment (aiming at health improvements), rehabilitation (aiming at improvements in functioning, and mitigation of the effects of disability), and palliation (aiming at improvements in quality of life). The salience of each of these activities varies across the disability cascade; promotion and prevention dominating for those yet to show declines in intrinsic capacity, treatment and rehabilitation for those with early isolated declines, and rehabilitation and palliation for those who are frail and dependent, some of whom will be nearing the end of life. There will be much blurring across these boundaries. As our data indicates, promotion and prevention are relevant throughout, yet not well covered in ICOPE; integration with the WHO Package of Essential Noncommunicable disease interventions (PEN) will be important since cardiovascular risk is high, and detection and control of hypertension and diabetes sub-optimal(34,37,38).

Community healthcare workers (CHWs) have an essential part to play if universal health coverage is to extend to older adults in low-resource settings. There has been interest in extending the roles of this cadre from maternal and child health, sanitation and infection control to include prevention and control of noncommunicable disease. There is early evidence of feasibility and effectiveness in cardiovascular risk reduction, detecting hypertension and diabetes(39,40), and in supporting the management of hypertension(41,42). Coordinating the integrated care of older people in the community would be a logical next step, capitalising on the CHWs unique outreach capacity and knowledge of older people and their families(43). Assessing intrinsic capacity in older adults does not require specialist skills, and can be done by CHWs in low-resource settings using a structured tool after brief training(43). Outreach may be necessary for early identification of DIC; in the current analysis, although those with DIC are slightly more likely to use healthcare services, this is mainly confined to those who are also frail or dependent.

Outreach will also help to engage this group who may otherwise find it difficult to access services. In an earlier report from the same cohort we have highlighted the inequity in access linked to poverty and lack of health insurance (44). Nevertheless, the challenges would be considerable. A fully-fledged system might comprise universal screening, followed by more detailed assessment for the two-thirds or more of the older population who could benefit from one or more ICOPE interventions. Optimal intervals of reassessment for screen negative and screen positive groups would need to be established. This would require a significant diversion of limited healthcare resources, and the cost-effectiveness of the investment is uncertain. Screening would not be warranted if resources and systems were not in place to provide the recommended interventions.

### Implications for research

The evidence used to support most of the ICOPE recommendations is indirect(44). Interventions were mostly evaluated in HIC using experienced senior nurses, physical or occupational therapists, or nutritionists often with specialist support, and full access to secondary care referral pathways. In LMIC, with few specialists, task-shifting to trained and supported non-specialist healthworkers will be a key feature of any accessible healthcare system for older people. This need not imply an attenuation of effect; in the previously cited systematic review, effect sizes were larger for earlier studies (pre-1993), before systematic approaches to increasing coverage of treatment and care for older people were generalised in HIC(17).

While many of the ICOPE interventions can be delivered at home by CHWs, this will not be sufficient; there will need to be clear and well-functioning referral pathways for, for example, clinical assessment and treatment, optometry and cataract surgery, and hearing aid services. Pilot intervention development will be needed for each of the recommendations to confirm feasibility and optimise modes of delivery in different contexts. ICOPE is a complex intervention, comprising a package of interventions, delivered flexibly to an individualised and person-centred care plan, involving different cadres and different levels of the health system, targeting the older person and their carers through direct physical intervention, behavioural and environmental change. Health systems research should focus on the planning, financing, resourcing and organisational change management that might be required to implement the ICOPE program at scale. Implementation science will be needed to understand the nature of the potential benefits, the underlying mechanisms, and how these can best be achieved. Ultimately, cluster randomised controlled trials generating evidence on cost-effectiveness are most likely to inform funding decisions. Most importantly, no opportunity should be lost to evaluate pilot projects, and to ‘learn by doing’.

## Conclusion

If the WHO’s ‘triple billion’ targets are to be achieved with equity, then it is essential that focused attention is given to closing the health access and outcome gaps experienced by older people, particularly those disadvantaged by disability, living in poverty, and in low-resourced areas. Implementation and scale up of the Integrated Care for Older People (ICOPE) program offers a conceptually simple, ‘bottom-up’ and public health orientated approach to ensuring that the benefits of universal health coverage and the enjoyment of better health and wellbeing are fairly distributed and inclusive. Effective implementation will require political will, prioritisation, investment, health system strengthening and restructuring. Research has a crucial role to play in guiding and evaluating initial attempts at implementation, and in providing the evidence that governments and policymakers require to prioritise action and mobilise investment.

## Data Availability

The data underlying this study are restricted, as participants did not consent to sharing their information publicly. Data are freely available from the 10/66 Dementia Research Group public data archive for researchers who meet the criteria for access to confidential data. Information on procedures to apply for access to data is available at https://www.alz.co.uk/1066/1066_public_archive_baseline.php, or by contacting Prof Martin Prince at.

## Authors’ contributions

MP wrote the manuscript and conducted the analyses. All of the authors worked collectively to develop the protocols and methods described in this paper. MP leads the 10/66 Dementia Research Group; JJLR (Cuba - Havana), AV (Cuba – Matanzas), DA (Dominican Republic), MG (Peru), AS (Venezuela), ALS (Mexico), KSJ (Vellore, India), ATJ (Chennai, India), and YH (China – assisted by ZL) are principal investigators responsible for the field work in their countries. All other authors reviewed the report and provided further contributions and suggestions. All authors read and approved the final report.

## Acknowledgments

Funded by Wellcome Trust (GR066133 – Prevalence phase in Cuba; GR080002-Incidence phase in Peru, Mexico, Cuba, Dominican Republic, Venezuela and China and data analysis across all centres) - http://www.wellcome.ac.uk/, World Health Organization (Prevalence phase in Dominican Republic and China) - www.who.int, US Alzheimer’s Association (IIRG–04–1286 - Prevalence phase in Peru, and Mexico) - http://www.alz.org/research/alzheimers_grants/, FONACIT/ CDCH/ UCV (data collection in Venezuela) - http://www.fonacit.gob.ve/, Puerto Rico Legislature (data collection in Puerto Rico), and the European Research Council (ongoing data collection, and further analyses of existing data - ERC-2013-ADG340755 LIFE2YEARS1066. The funders had no role in study design, data collection and analysis, decision to publish, or preparation of the manuscript. MJP, King’s College London is funded by the National Institute of Health Research (NIHR) Global Health Research Unit on Health System Strengthening in Sub-Saharan Africa, King’s College London (GHRU 16/136/54) using UK aid from the UK Government to support global health research. The views expressed in this publication are those of the author(s) and not necessarily those of the NIHR or the Department of Health and Social Care.

